# Incorporating Genetics Services into Adult Kidney Disease Care

**DOI:** 10.1101/2022.05.26.22275530

**Authors:** Kelsie Bogyo, Natalie Vena, Halie May, Hila Milo Rasouly, Maddalena Marasa, Simone Sanna-Cherchi, Krzysztof Kiryluk, Jordan Nestor, Ali Gharavi

## Abstract

Studies have shown that 1 in 10 adults with chronic kidney disease has a genetic component to their disease. However, genetic services in adult nephrology are limited. An adult Kidney Genetics Clinic was established within the nephrology division at a large urban academic medical center to increase access to genetic services and testing in adults with kidney disease. Between June 2019 through December 2021, a total of 363 patients were referred to the adult Kidney Genetics Clinic. Of those who completed genetic testing, a positive, diagnostic finding was identified in 27.14% and a candidate diagnostic finding was identified in 6.67% of patients, while a non-diagnostic positive finding was identified in an additional 8.57% of patients, resulting in an overall yield of 42.38% for clinically relevant genetic findings in tested patients. A genetic diagnosis had implications for medical management, family member testing, and eligibility for clinical trials. With the utilization of telemedicine, genetic services reached a diverse geographic and patient population. Genetic education efforts were integral to the clinic’s success, as they increased visibility and helped providers identify appropriate referrals. Ongoing access to genomic services will remain a fundamental component of patient care in adults with kidney disease.

## Introduction

Chronic kidney disease (CKD) is a debilitating disorder associated with significant morbidity and mortality and affects approximately 15% of adults in the United States (CDC, 2021). A diagnosis of CKD is often based on overlapping, non-specific clinical features, and histology findings, and in adults, the underlying etiology remains unknown in many cases. While the underlying genetic contribution to pediatric kidney disease is well established, the role of genetics in adult-onset kidney disease has been overlooked until recently.

Recent studies have shown that 1 in 10 adults with CKD have a genetic component to their disease (Groopman E E, 2019). This study, along with several others (Vivante A, 2016; Connaughton DM, 2019), identified monogenic etiologies of kidney disease across the spectrum of clinical diagnoses in kidney disease. Notably, a diagnostic yield of ∼20% was reported in those with nephropathy of unknown origin (Groopman E E, 2019; Hays T, 2020). Similarly, a high yield for genetic diagnosis has been found in individuals with cystic kidney disease or congenital anomalies of the kidney and urinary tract (CAKUT). Genetic causes of other types of kidney disease, such as glomerulopathies, tubulopathies, and complement disorders, are also common, especially when accompanied by other risk factors for a genetic etiology, such as a family history of kidney disease, young age of onset, or extra-renal features (Cocchi E, 2020). Thus, genetic testing is an emerging tool to aid in identifying the underlying etiology and diagnosis of CKD.

Establishing a genetic diagnosis in a patient with kidney disease can clarify the diagnosis, identify extra-renal manifestations for which the patient may be at risk, and clarify the genetic risk to other family members. In addition, a genetic diagnosis can also aid in the identification of eligible donors for kidney transplantation (Garg AX, 2020; Niaudet, 2010; Mann N, 2019), prevent the use of unnecessary immunosuppression (Preston R, 2019), and increase patient eligibility for a growing number of clinical trials based on genetic disease status (Milo Rasouly H, 2018). As such, clinical genetic testing is increasingly being recommended as a tool for the diagnosis and management of kidney disease (Knoers N, 2022).

With these emerging data, the nephrology medical specialty is increasingly utilizing genomic medicine in clinical practice, similar to other specialties such as oncology (Hampel H, 2015) or cardiology (Hershberger R E, 2018; Mital S, 2016), where genetics clinics have become well integrated into clinical practices and are associated with positive outcomes, such as increased knowledge and positive health behaviors, and decreased anxiety and decisional conflict (Madlensky L, 2017). Newly formed renal genetic clinics have reported their early experiences with genetic testing (Amlie-Wolf L, 2021; Thomas C P, 2020; Mallett A, 2016; Alkanderi S, 2017; Pode-Shakked B, 2022; Lundquist A L, 2020; Elhassan EAE, 2022), utilizing the expertise of clinical geneticists, genetic counselors, and nephrologists to guide the interpretation, clinical assessment, and correlation of genetic findings. The integration of genetic counselors in nephrology aims to address nephrologists’ lack of training and low confidence in interpreting genetic results (Jayasinghe K, 2020; Berns, 2010). However, utilization of genetic services remains limited in the adult nephrology population and these recent renal genetics clinics have reported diverse initial experiences in terms of clinical setting, roles of members of the team and population served. Here, we present the experience establishing and maintaining a kidney genetics clinic serving a relatively high patient volume within an adult nephrology division at a large, urban academic medical center.

## Methods

### Laying the Foundation

Prior to establishing the Kidney Genetics Clinic, several key research initiatives were implemented within the Division of Nephrology at Columbia University Irving Medical Center (CUIMC) which were central to the success of the clinic. Following the study on 2,187 patients enrolled at CUIMC that reported that 1 in 10 adults with CKD has a monogenic form of kidney disease (Groopman E E, 2019), a pilot study was carried out to return those research genetic results to participants (Nestor JG, 2020). In a continuation of these studies, a monthly genetic variant sign-out meeting and educational series were established. At the sign-out meetings, which involved nephrologists, geneticists, genetic counselors, and genetics researchers, clinical and genetic information on research cases from the ongoing research studies were discussed and the group agreed upon which variants would be clinically confirmed and returned to the participants. The referring clinical nephrologists were involved throughout this process. The educational series included an interactive, biweekly renal genetics case series to familiarize and engage with the nephrologists within the Division of Nephrology on renal genetics. Topics from these cases highlighted subjects such as when to suspect a genetic kidney condition, genetic test selection considerations, aspects and interpretation of genetic results, key management implications of the genetic diagnosis, and cascade testing of at-risk family members. Basic genetic topics and vocabulary such as the types of inheritance patterns, penetrance, and variable expressivity, as well as their clinical implications were introduced and discussed. Familiarizing nephrologists to the research workflow and educating them on genetic topics commonly encountered in the clinic, facilitated the establishment and utilization of the Kidney Genetics Clinic.

### Setting and Clinic Structure

In June 2019, an adult Kidney Genetics Clinic was created within the Division of Nephrology in the Department of Medicine at CUIMC. CUIMC is a large academic and clinical medical institution located in the Washington Heights neighborhood of Manhattan, NY.

Within the Kidney Genetics Clinic, two visit types were available for new patients: *Full Genetic Consults* (staffed by a genetic counselor (GC) and nephrologist) and *Genetic Counseling Visits* (staffed by a GC only). *Full Genetic Consults* involve obtaining a complete medical and family history, performing a personalized genetic risk assessment, and when applicable, obtaining informed consent and sample coordination for clinical genetic testing. *Genetic Counseling Visits* typically involve obtaining informed consent and sample collection for a specific genetic test recommended by the patient’s treating nephrologist, obtaining consent for cascade testing of a known familial variant, or counseling on genetic test results. To return results and follow-up on genetic testing previously ordered by the Kidney Genetics Clinic, the clinic offers *Return Patient Visits* (staffed by a GC only). The full workflow for the Kidney Genetics Clinic from referral to return of results can be found in Figure 1.

**Figure 1.**
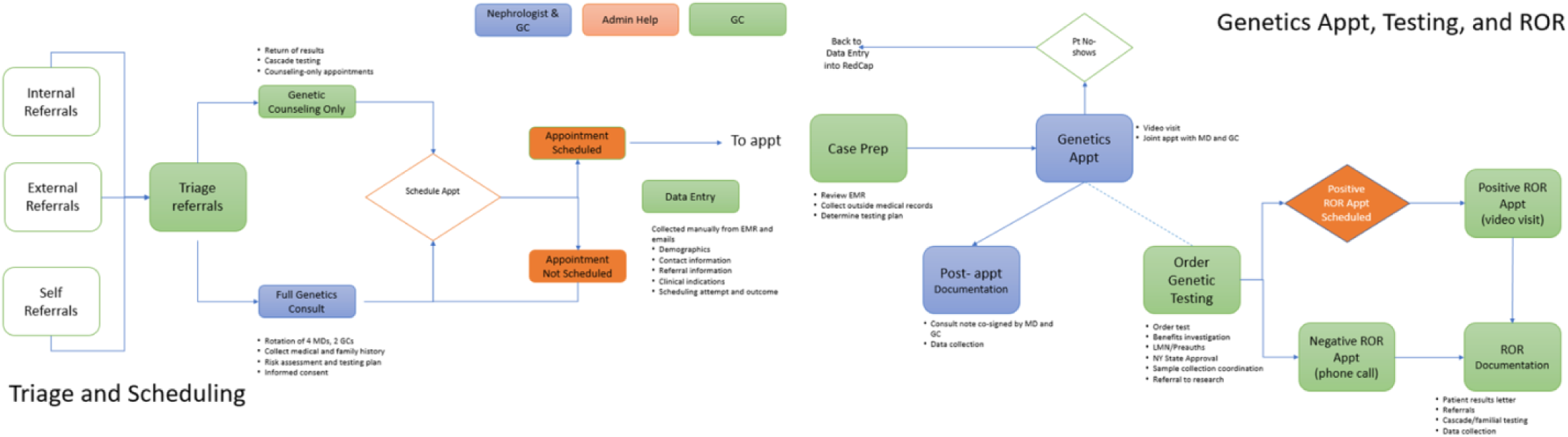
Current workflow for referrals received in the Kidney Genetics Clinic through return of genetic results at Columbia University Irving Medical Center.

The clinic was held weekly with a maximum of 2 concurrent visits and six visits a day (six 1-hour appointment slots for Full Genetic Consults and six 1-hour Genetic Counseling Visits or Return Patient Visits). Each clinic was staffed by two genetic counselors and one of four rotating nephrologists with specialized training or interest in genetics and precision medicine. Administrative help was also provided for scheduling and insurance support.

Both Full Genetic Consults and Genetic Counseling Visits occurred in-person at the Washington Heights medical campus location prior to March 2020, at which point the clinic briefly halted operations until May 2020, then transitioned almost exclusively to telemedicine (virtual video appointments) due to the COVID-19 pandemic. Thereafter, virtual video appointments became the default method for all new patient genetic consults, although in-person appointments remained available based on patient preference. Return Patient Visits for return of results appointments occurred in-person, by phone, or video prior to the pandemic, and transitioned exclusively to telemedicine format in March 2020.

### Referrals

Providers were given many options regarding how to refer their patients, including messaging a clinic-specific email, Electronic Medical Record (EMR) referral, online referral form, phone call, or email to a clinic staff’s personal email. The phone number, email, and online referral were accessible online (Supplemental Figure 1). Referrals were accepted from providers as well as directly from patients. All referrals and accompanying clinical information were stored in a Columbia University REDCap project. The administrative staff made two contact attempts to schedule each referred patient and all outcomes were documented.

### Genetic Testing

All genetic tests ordered through the Kidney Genetics Clinic were sent out to commercial CLIA (Clinical Laboratory Improvement Amendments)-certified laboratories and variants were classified according to the laboratory protocols. A positive diagnostic finding was considered when a pathogenic or likely pathogenic variant(s) was identified in a gene that fully or partially explained the patient’s features. A positive non-diagnostic finding was either a common risk factor (such as a high risk *APOL1* genotype) or a pathogenic or likely pathogenic variant which did not explain the patient’s kidney disease. A candidate diagnostic finding was defined when there was clinical suspicion of a variant of uncertain significance being causative because the gene was associated with a condition that had a significant clinical overlap with the patient’s presentation.

### Data analysis

A retrospective chart review was performed on all patients referred to the Kidney Genetics Clinic from June 2019 to 2021. Demographic information, clinical features, family history, referral indications, and contact outcomes were collected from referral documents and data. For patients scheduled in the Kidney Genetics Clinic, demographic and referral information, detailed clinical and family history, genetic testing results, as well as management implications and referrals were collected from the EMR, testing documents, and directly from the patients.

All data collection was documented in the REDCap database and performed in accordance with the Genetic Studies of Constitutional Disorders protocol, (Institutional Review Board Protocol Number AAAS7948) approved by the Columbia University Institutional Review Board.

## Results

### Referral Indications and Methodologies

Between June 2019 and December 2021, a total of 363 patients were referred to the Kidney Genetics Clinic from multiple sources, including internal CUIMC providers, external providers, and self-referrals (Figure 2). Referrals from CUIMC providers (internal referrals) represented 73% of referrals, and spanned across 8 different divisions and departments, including Nephrology, Cardiology, Cancer, Genetic, Reprogenetics/OBGYN, Ophthalmology, Internal Medicine, and Gastroenterology. Eleven percent of referrals were self-referrals, and 16% of referrals came from over 30 different external institutions (Supplemental Table 1). 76.3% of referrals were received directly through the Kidney Genetics Clinic email, 6.67% through the EMR, 4.1% through the clinic online referral form, and 13.0% were received through other mechanisms, such as a phone call or personal correspondence (Supplemental Table 1). After the initial transition to the Epic EMR software, EMR referrals became a more popular referral method for the clinic. While the email address was still most often used by referrals from external institutions, online referral forms were more commonly used in this group of referrals (Supplementary Table 1). Finally, patients who self-referred were most likely to do this by contacting the phone number or by sending a personal email.

**Figure 2.**
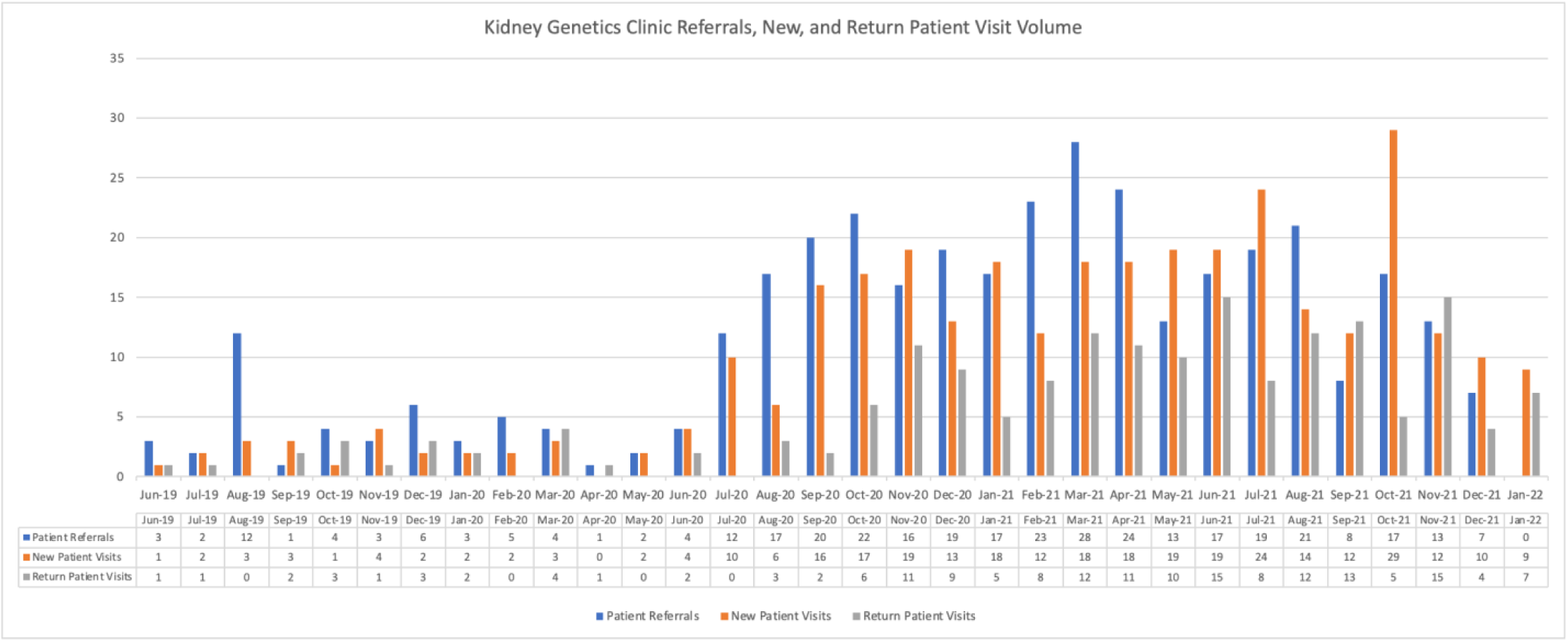
Kidney Genetics Clinic volume of referrals, new patient consults, and return patient visits from June 2019 to January 2022.

Of all patients referred, 304 patients (84%) were scheduled for a new patient genetic consult. The average wait time between initial referral request and scheduled appointment was of 37 days. For patients referred for donor evaluation, the average wait time from referral to appointment was 5 days. There was no significant difference in scheduling rates between internal, external, or self-referrals (Supplemental Table 1). Among the referrals where a new patient genetic consult was not scheduled, no contact with the patient (n=33) was the most cited barrier, followed by patient declined or lack of interest (n=15). Access limitations (n=7), including lack of professional state licensure, technology issues, and cost, were rare but reported barriers to scheduling new patient genetic consults. Four patients were also referred to the Kidney Genetics Clinic, but their referral was determined to be not clinically relevant for the Kidney Genetics Clinic and they were referred elsewhere.

Overall, 304 patients were scheduled. Of those, who did not present to their initial visit, 20 were re-scheduled for a later date. Ultimately, 324 new patient genetic consults were scheduled in the Kidney Genetics Clinic at CUIMC, and 279 patient visits were completed, resulting in an overall no-show rate of 13.9% (Figure 2). In addition to new patient genetic consults, 176 follow-up visits occurred from June 2019 to January 2022. Overall, most scheduled visits were telemedicine appointments via virtual video visits (61%), or phone appointments (33%, Table 1).

**Table 1.** Scheduled Appointments by Visit Type and Modality in the Kidney Genetics Clinic

| Visit Type | In-person | Phone | Video | Total Visits |
| --- | --- | --- | --- | --- |
| Complete Genetics Consult | 28 | 3 | 233 | 264 |
| Genetic Counseling Visit | 2 | 20 | 38 | 60 |
| Return Patient Visit | 1 | 143 | 32 | 176 |
| Total | 31 (6.2%) | 166 (33.2%) | 303 (60.6%) | 500 |

Most patients seen in the Kidney Genetics Clinic were referred to clarify a clinical diagnosis (n=186), but referral indications also included clarification of biopsy finding (n=20), transplant donor and recipient evaluation (n=53), and cascade and family member genetic testing (n=39). Patients had a variety of kidney-related clinical indications and diagnoses, many of which were overlapping. These included: suspected or clinical diagnosis of a collagenopathy (n=49), hematuria and/or proteinuria (n=45), focal segmental glomerulosclerosis (FSGS) (n=37), cystic kidney disease (including PKD) (n=35), tubulopathy or electrolyte disorder (n=28), tubulointerstitial disease (n=26), complement dysregulation (n=12), CAKUT (n=8), tumor or cancer (n=5), and CKD of unknown etiology (n=29). In addition, 32 patients were healthy or unaffected individuals.

### Patient Demographics

The Kidney Genetics Clinic patient population was relatively diverse, with 49% who self-identified as white, 21% Hispanic/Latinx, 15% Black/African American, 7% Asian, 1% Native Hawaiian/Pacific Islander, and 7% other or preferred not to specify (Table 2a-e). Most consultations were in English, but 5.7% were held in Spanish. The majority were female (57%), and of those seen in the Kidney Genetics Clinic, most (59%) patients had private insurance, while 28% had government insurance (Medicare or Medicaid), and insurance type was unknown for 13%. The average age of the patient population was 44 years old and ranged from 18 to 87 years. A family history of kidney disease was reported in 104 patients (37%) and 69 patients (25%) had a personal or family history of a known genetic diagnosis at the time of their appointment (Table 3a-c). The Kidney Genetics Clinic patient population largely resided in the NY tri-state area (87%), but 13% resided in ten additional states, three other countries, and one US territory.

**Table 2.**
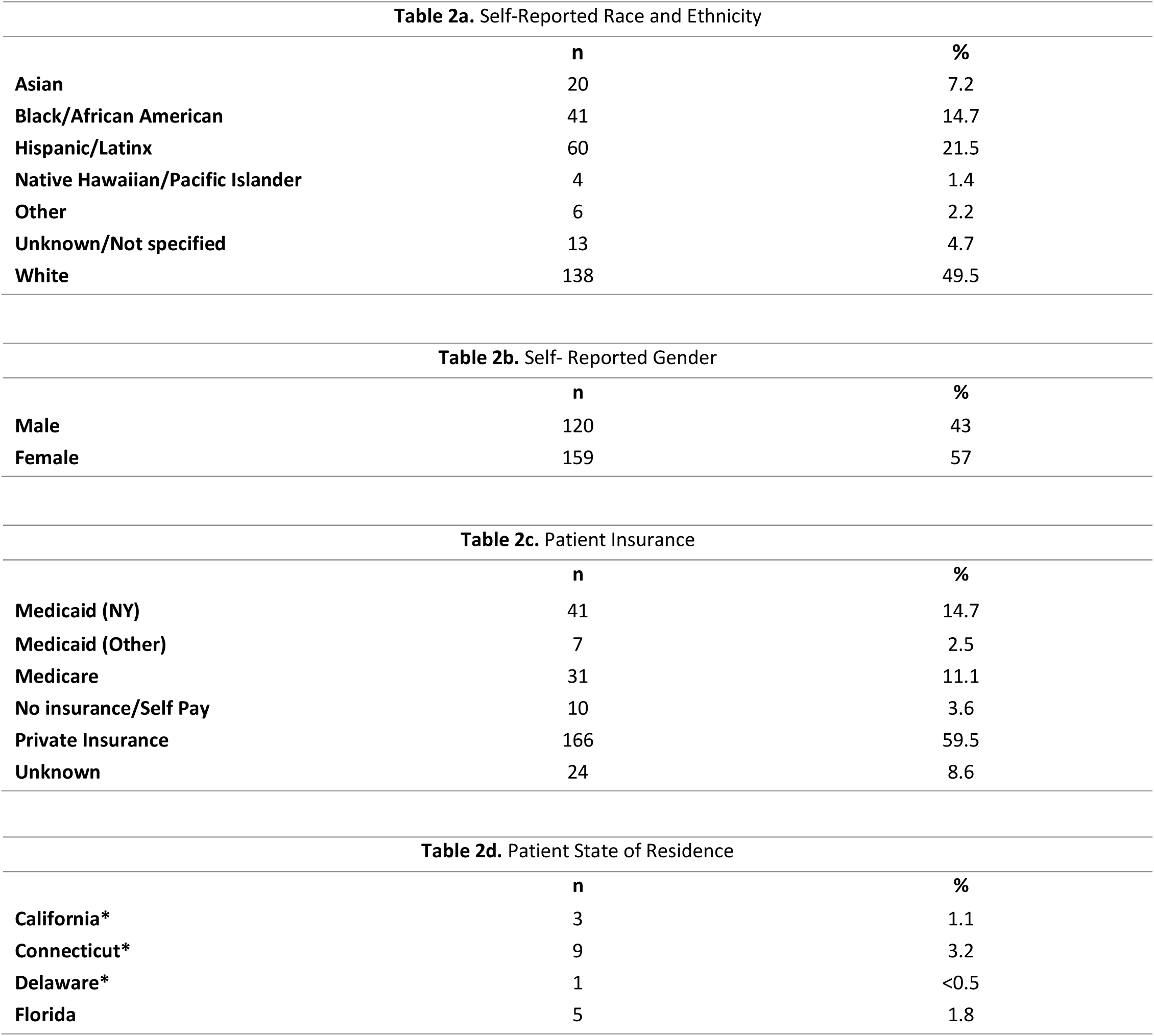
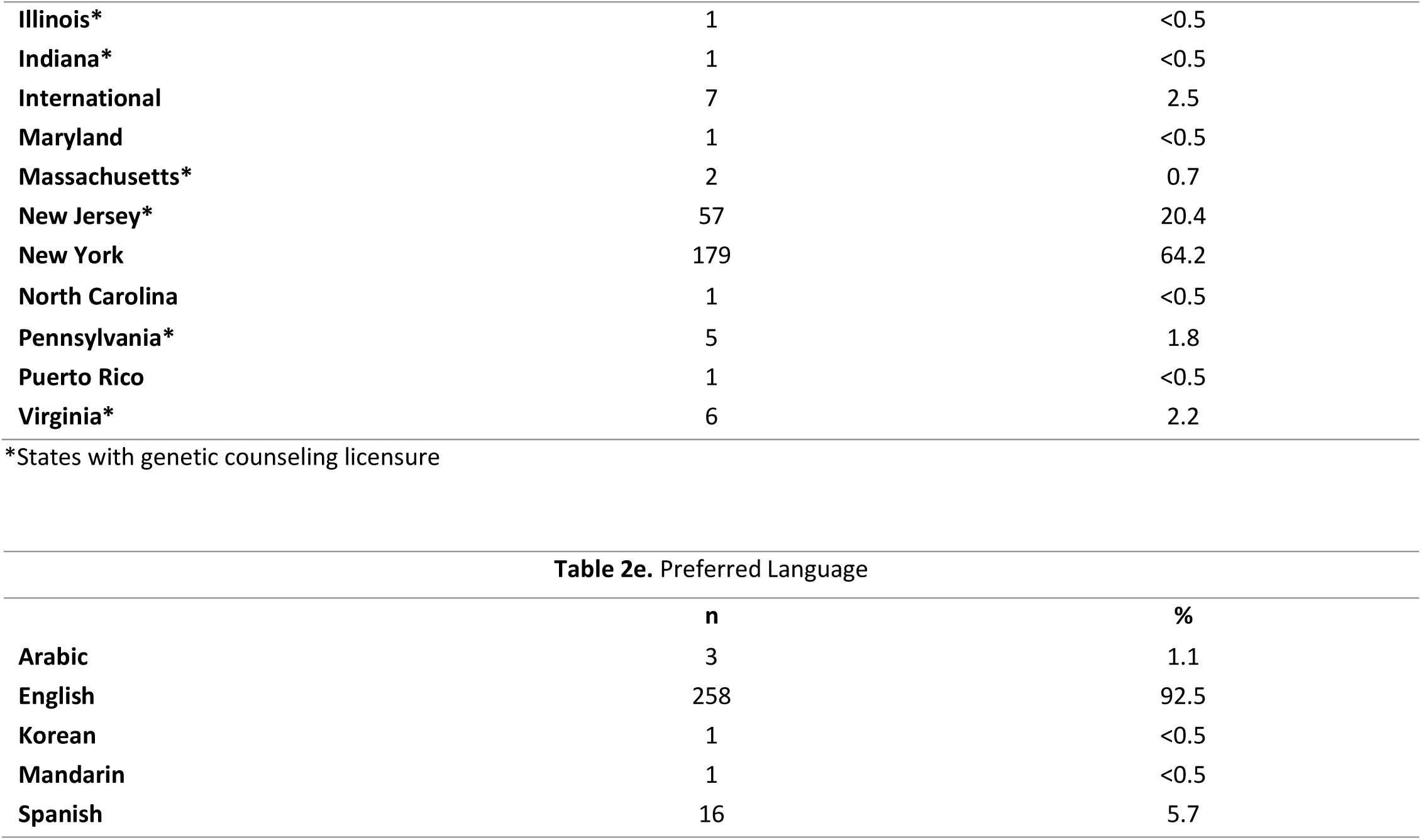
Demographic details of the Kidney Genetics Clinic Patient Population

**Table 3.**
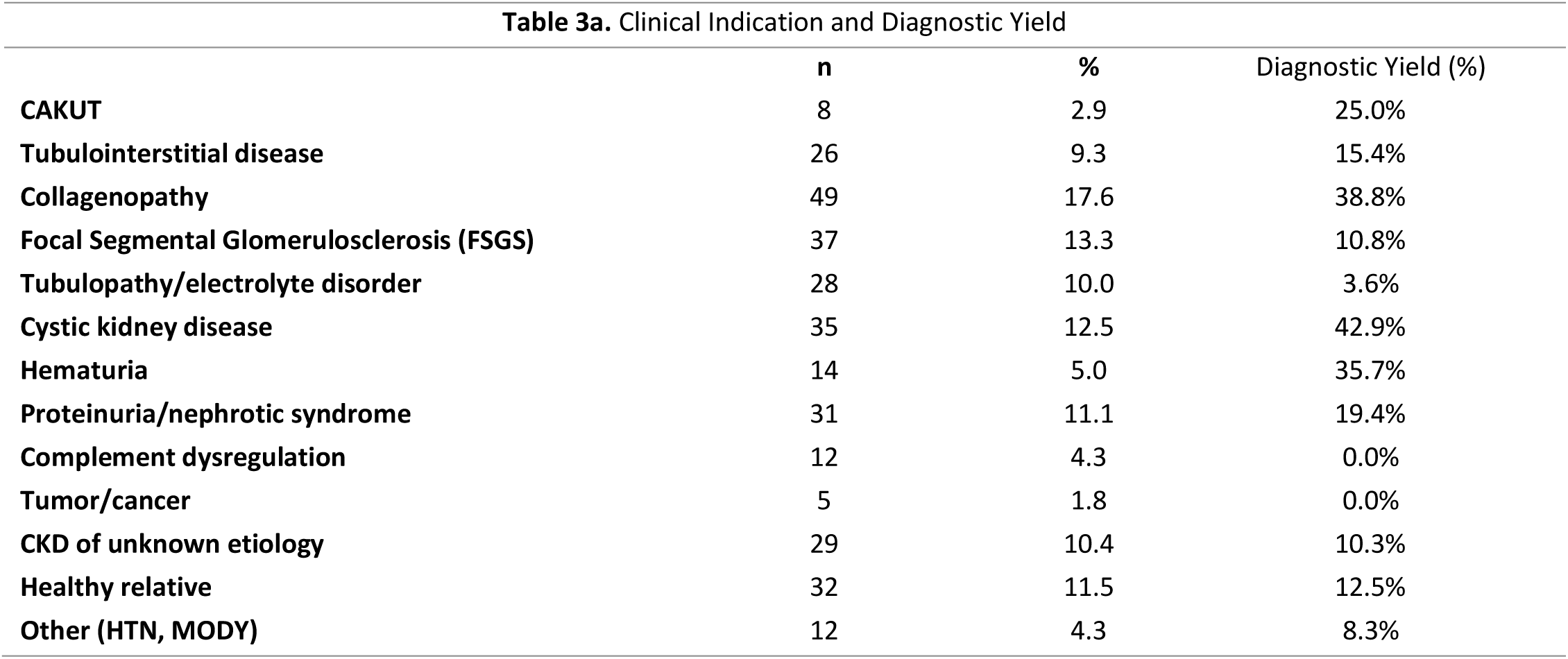
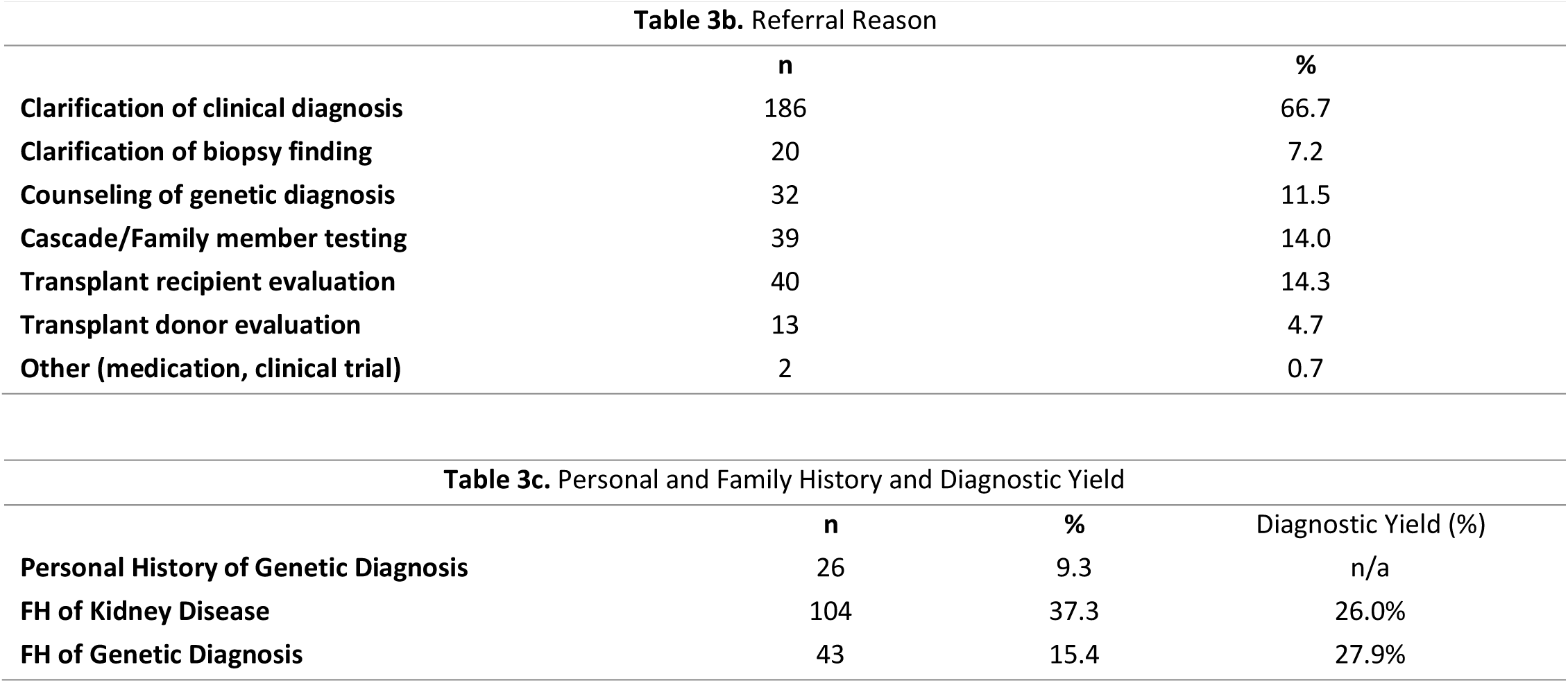
Referral and Clinical Information of the Kidney Genetics Clinic Patient Population

### Genetic Test Ordering and Results

As part of the new patient consults, several commercial CLIA-certified genetic tests were ordered, including small (<100 genes) and large (>100 genes) multi-gene panels, exome sequencing, chromosome microarrays, single-gene sequencing, and targeted variant testing (Supplemental Table 2). A total of 249 clinical genetic tests were ordered on 82.8% (n=231) patients, with 18 patients having concurrent or reflex genetic testing ordered. Nineteen tests were canceled, as the patients never submitted samples for testing, and results were still pending for five tests at the time of publication. In patients for whom genetic testing was not ordered, reasons included: testing not clinically indicated (n=6), patient declined testing or did not provide consent (n=15), genetic testing results already available (n=24), and financial concerns (n=3).

Presently, 225 genetic testing results were complete and available for 210 patients seen in the Kidney Genetics Clinic. Among these patients, a diagnostic finding was identified in 27.14% of patients and a candidate diagnostic finding was identified in 6.67% of patients, resulting in a genetic finding potentially explaining the patient’s kidney disease in 33.81% of the Kidney Genetics Clinic patient population. A non-diagnostic positive finding was identified in an additional 8.57% of patients. Non-diagnostic findings included secondary findings (n=1), carrier status in phase testing (n=5), and risk factors, such as an *APOL1* high-risk genotype (n=12).

For results considered positive (diagnostic or non-diagnostic) or candidate diagnostic findings, 98 variants in 28 unique genes were identified (Figure 3c, Supplementary Table 3). One patient was found to have variants in two genes associated with Alport syndrome (variants in *COL4A4* and *COL4A5*), two patients were found to have dual diagnostic findings, and four patients were found to have both a (Elhassan EAE, 2022)diagnostic or candidate diagnostic finding in addition to a non-diagnostic finding. For two patients, the original results were classified as candidate diagnoses, but since the initial report was issued, the results were upgraded to diagnostic findings by the laboratory. This process was facilitated by our input to the laboratories. The diagnostic yield was found to be highest among patients with either cystic kidney disease or clinical suspicion of a type IVa collagenopathy/Alport syndrome based on either clinical features or kidney biopsy results (Figure 3b, Table 3a).

**Figure 3.**
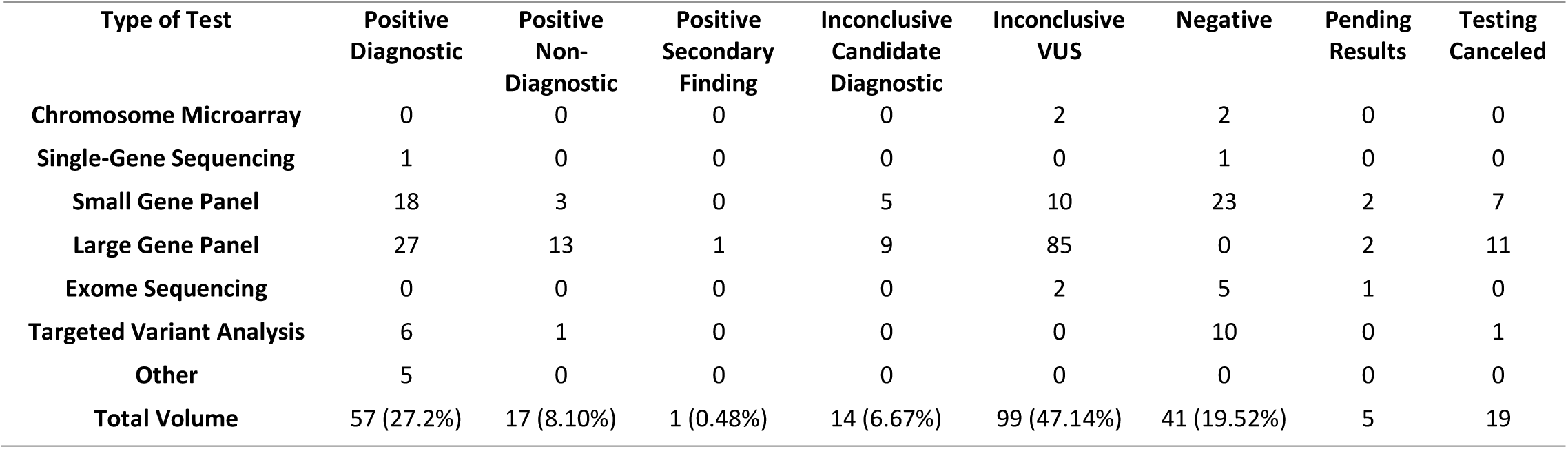
Genetic Test Results.

**Figure 3b.**
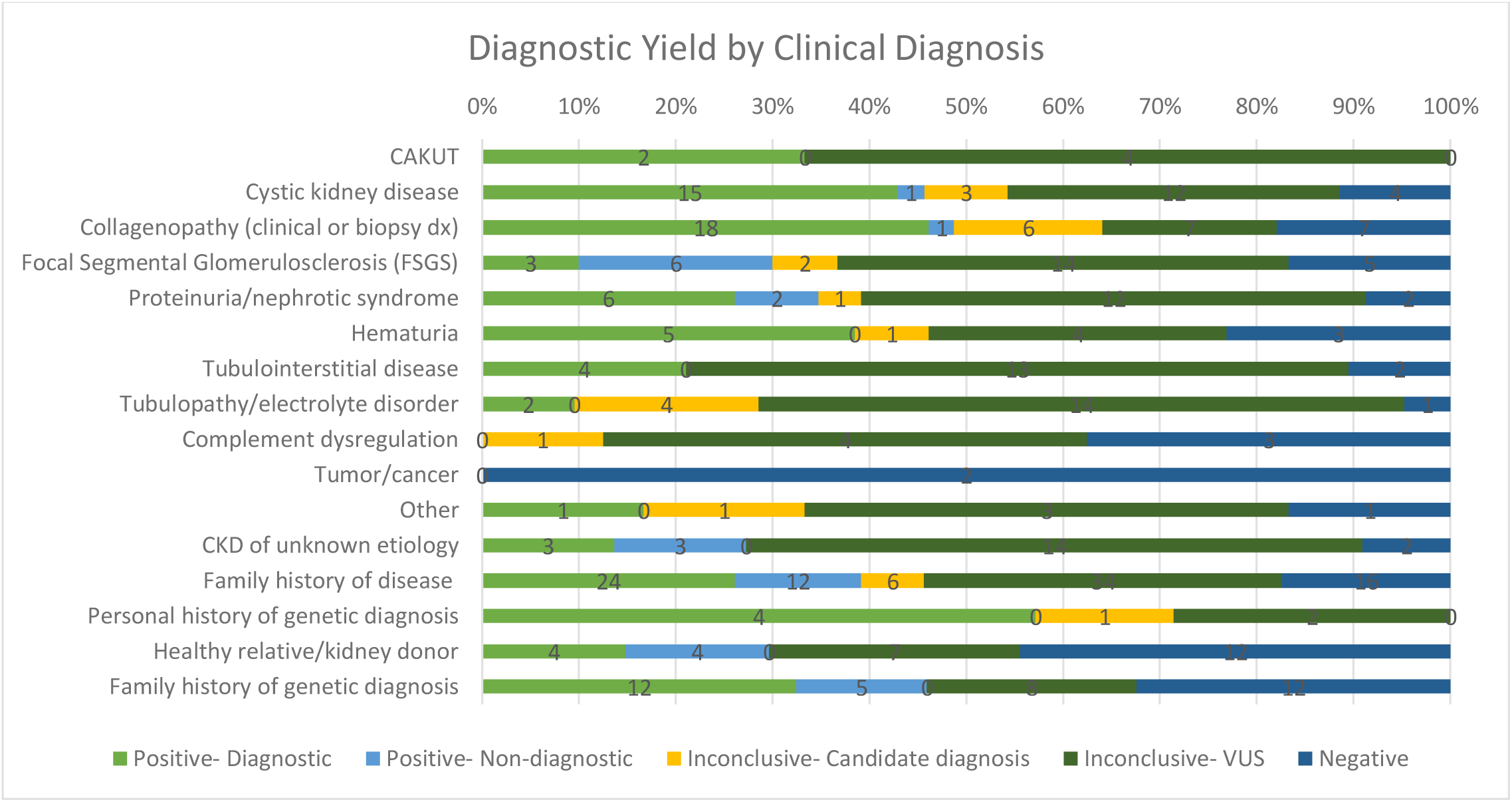
Diagnostic Yield by Clinical Indication. The highest diagnostic yield was found among patients with either cystic kidney disease or clinical suspicion of a type IVa collagenopathy/Alport syndrome based on either clinical features or kidney biopsy results.

**Figure 3c.**
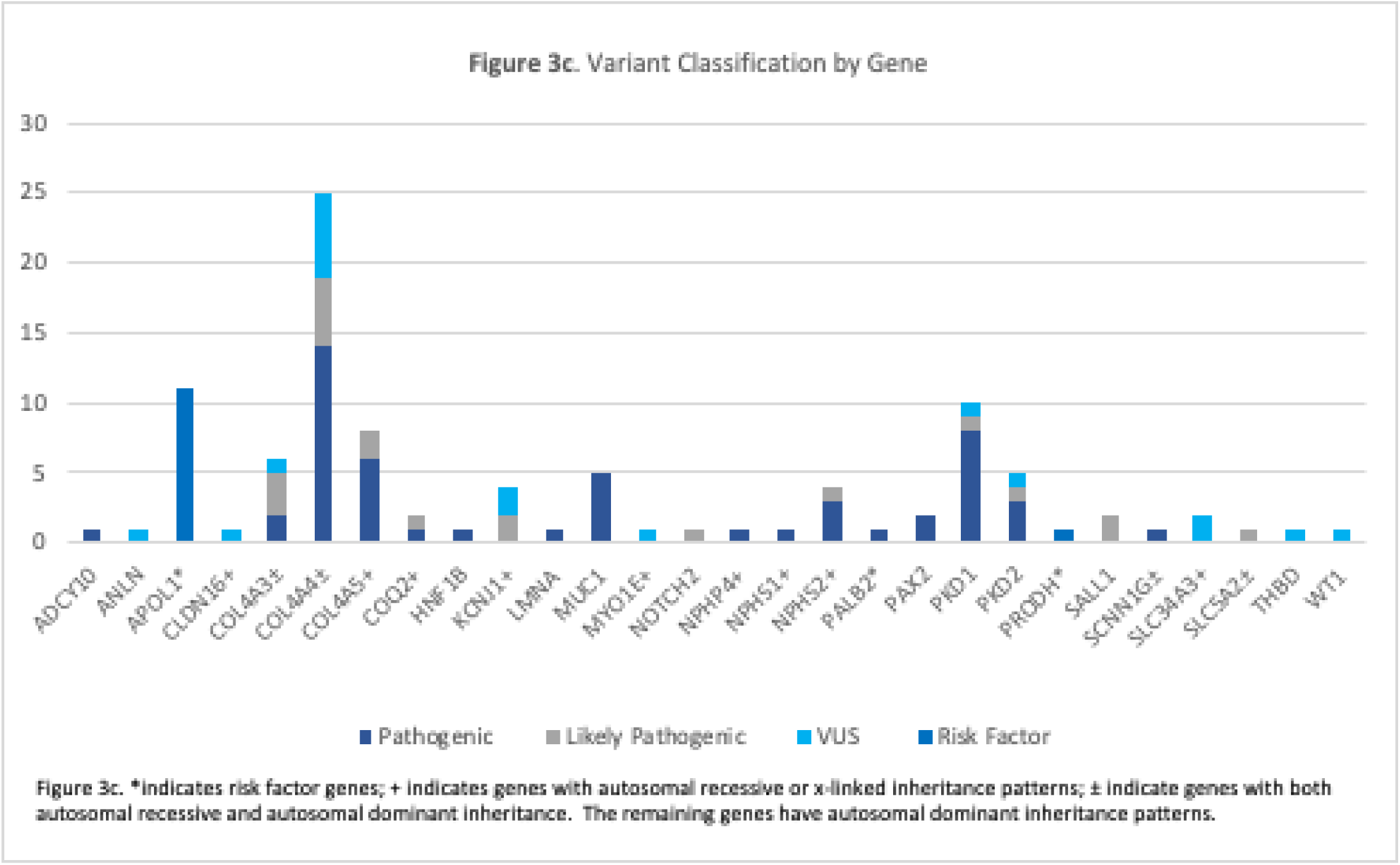
Variant Classification by Gene

### Impact on Management

Based on their genetic results, a referral to at least one specialist outside of nephrology was made for 40 patients (including ENT, ophthalmology, cardiology, hepatology, etc.). Of the 53 patients referred for transplant evaluation, 15 were found to have a genetic finding that could impact donor selection, and family member testing was recommended in 12 patients for VUS resolution. In two patients, a genetic diagnosis of ADPKD was established and based on this, they were able to obtain insurance coverage of tolvaptan. In addition, 28 patients with at least one pathogenic or likely pathogenic in *COL4A3, COL4A4* or *COL4A5* were potentially eligible for participation in ongoing clinical trials for treatment of Alport syndrome. If they met other criteria for enrolling, they were referred to the trial. Patients with non-diagnostic findings were referred to ongoing research studies at the medical center to further elucidate the genetics of kidney disease.

## Discussion

Genomic medicine has entered mainstream medicine and is increasingly available in medical specialties. However, integration of genomic medicine in nephrology has been limited, in part, by nephrologists’ lack of confidence and knowledge of genetic information (Jayasinghe K, 2020; Berns, 2010). To address this, nephrologists and genetic counselors have collaborated to create a comprehensive Kidney Genetics Clinic within the Division of Nephrology at CUIMC. This clinic provides complete genetic care for adult patients with kidney and associated diseases.

### Successes

The structure of the Kidney Genetics Clinic was created such that it provides flexibility to providers, offering multiple appointment types and points of entry into the clinic workflow. This workflow was specifically designed to cater and appeal to the diverse level of providers’ genomic literacy. If, for example, the referring provider had already made a clinical diagnosis of a genetic condition and had a clear and targeted idea of what should be tested for, the patient was scheduled for a Genetic Counseling Visit, in which the patient was seen for a more targeted counseling session and to consent for the most appropriate test. The Genetic Counseling Visits provided access to limited genomic services while integrating care with patients’ treating doctors. However, if the patient was suspected to have an underlying genetic condition based on their presentation, but a specific gene or condition was not highly suspected, the patient was seen for a Full Genetic Consult with a genetic counselor and nephrologist.

The efficiency of the Kidney Genetics Clinic is highlighted by the short wait time for new patient visits, with the average time from referral to appointment being 37 calendar days, and 5 days for patients referred for donor evaluation. This wait time is likely an overestimate of the typical patient’s wait time to an appointment, as this includes those who were contacted multiple times over the course of a few weeks as well as patients who were never able to be contacted and were re-referred months later. Comparatively, the average wait time for a new patient visit in a traditional genetics clinic is on average 3-4 months (Maiese DR, 2019). In the nephrology setting, especially in the setting of transplantation, there can be an urgency associated with these genetic results and the timeline becomes critical. This model has the capacity to reduce barriers to genetic testing and counseling such as long wait times for general genetics clinics.

While data is not available on the direct cost incurred by patients for the tests, the high uptake of genetic services, as well as low test cancellation rates due to financial constraints, suggest that cost is not a significant barrier to accessing genomic services in nephrology. While the most common reason for test cancelation was due to patient non-compliance (i.e. not sending in a sample or completing test consents), these rates were similar regardless of insurance status or type. This suggests that, although financial concerns cannot be ruled out as a consideration, it does not disproportionally affect a particular demographic in the Kidney Genetics Clinic.

### Telemedicine Utilization

The Kidney Genetics Clinic was established in June 2019, prior to the COVID-19 Pandemic but operations were significantly impacted in response to the global pandemic. In mid-March 2020, the clinic stopped all activities in order to divert clinical resources to the pandemic. The clinic slowly began to see new patients again in May 2020 as it switched to a telemedicine model, and all patient visits at this time occurred via video visit or phone call, depending on the needs of the patient. The relaxed medical professional licensure rules during this time greatly increased nephrologist and genetic counselors’ ability to provide virtual healthcare to patients across the country, significantly expanding access to genomic services in nephrology.

As such, the Kidney Genetics Clinic experienced a significant jump in number of referrals during the summer of 2020 (Figure 2), and the number of monthly referrals has since remained stable. Since then, telemedicine models have become much more prevalent, mostly motivated by the increased access to care that telemedicine can provide (Garfan S, 2021). There were no differences in conversion rate from referral to attending scheduled visits based on referral source or patient demographics, suggesting this model was successful in bringing genomic medicine to a diverse group of adults with kidney disease. By continuing to utilize a combination of in-person appointments and telemedicine, the Kidney Genetics Clinic will help genetic services reach a broad geographic region and diverse patient population.

While telemedicine has been reported to reduce barriers, barriers with this model still exist. For video visits, a smartphone and reliable internet access are required. Additionally, a certain level of technology literacy is required to join the visits. To accommodate these barriers, visits were conducted over the phone via conference call when the patient was unable to join the video visit. The built-in telemedicine capability in the EMR also facilitated patient communication.

Given the success of the implementation of telemedicine in this clinic, the Kidney Genetics Clinic continued with a primarily telemedicine model even after routine in-person visits began to resume in the nephrology division.

Telemedicine remained a preferred method of visit for many patients because of the convenience, as well as the high proportion of nephrology patients who are immunocompromised. However, if a patient requested an in-person visit or if an in-person visit was deemed more accessible by the patient and provider, based on technological issues or language barriers, in-person visits were available and accommodated on a case-by-case basis.

### Implications of Genetic Diagnoses

The identification of genetic diagnoses in this patient population has several clinical implications, including changes in management, eligibility for genetically stratified clinical trials, and treatment implications. The diagnoses in patients impacted several areas of clinical care, including referrals to specialists, kidney donor selection, clinical trial eligibility (for example, in patients with a genetic diagnoses of Alport Syndrome) and increased access to medications (such as tolvaptan in patients with *PKD1* variants). Those with non-diagnostic findings were referred to ongoing genetics and clinical research studies at the medical center to further elucidate the genetics of kidney disease.

### Future Considerations

The goal of the Kidney Genetics Clinic continues to revolve around increasing access to genetic testing and counseling and reducing barriers that might contribute to health inequities.

Since the onset of the COVID-19 pandemic, many states issued temporary emergency waiver of licensure requirements. This allowed for the clinic’s rapid expansion to states across the country. However, most of those waivers have now expired, and physician and genetic counselor licensure needs to be a consideration in seeing patients located in other states. Efforts in this area have involved obtaining both physician and genetic counselor licensure in the tri-state area (NY, NJ, CT) as well as in other states where the clinic receives a substantial number of referrals. However, the resource-intensive and time-consuming nature of applying for out of state licensure cannot be underestimated.

While educating nephrologists on the impact and importance of genetic counseling and testing contributed to the success of the clinic, there are still areas where additional physician education can improve referrals. Regular seminars and educational initiatives aimed at nephrologists and other advanced care practitioners in nephrology will continue to be integral. Additionally, offering more advanced day-long courses for specialists who are interested in starting to order genetic testing as part of their clinical practice may be beneficial. Expanding these educational initiatives to target other specialists who, though maybe not focused on kidney disease, might be able to notice indications that should that trigger a referral to genetics could also be beneficial. Finally, though the clinic offered the ability for patients to self-refer, few of the referrals came directly from patients. Future educational initiatives that target the general kidney disease patient population might raise awareness among patients about when to discuss genetic testing with their nephrology provider or refer one-self to genetic counseling.

## Data Availability

All data produced in the present study are available upon reasonable request to the authors

